# Placental transfer dynamics and durability of maternal COVID-19 vaccine-induced antibodies in infants

**DOI:** 10.1101/2023.12.08.23299716

**Authors:** Paola A. Lopez, Nadège Nziza, Tina Chen, Lydia L. Shook, Madeleine D. Burns, Stepan Demidkin, Olyvia Jasset, Babatunde Akinwunmi, Lael M. Yonker, Kathryn J. Gray, Michal A. Elovitz, Douglas A. Lauffenburger, Boris D. Julg, Andrea G. Edlow

## Abstract

Completion of a COVID-19 vaccination series during pregnancy effectively reduces COVID-19 hospitalization among infants less than 6 months of age. Elucidating the dynamics of transplacental transfer of maternal vaccine-induced antibodies, and their persistence in infants at 2, 6, 9 and 12 months, has implications for new vaccine development and timing of vaccine administration in pregnancy to optimize protection of the mother-infant dyad. We evaluated anti-COVID antibody IgG subclass, Fc-receptor binding profile, and activity against wild-type Spike and RBD, and five variants of concern (VOCs) in 153 serum samples from 100 unique infants. Maternal IgG1 and IgG3 responses persisted in 2- and 6-month infants to a greater extent than the other IgG subclasses, with highest persistence of antibodies that bind placental neonatal Fc-receptor as well as FcγR3A. Timing of maternal vaccination and fetal sex were drivers of antibody persistence in infants. Lowest persistence at 2 and 6 months was observed against the Omicron RBD-specific region. Maternal vaccine timing, placental Fc-receptor binding capabilities, antibody subclass, fetal sex, and VOC all impact the persistence of maternal vaccine-induced antibodies in infants up to 12 months.

## INTRODUCTION

The U.S. Centers for Disease Control and Prevention (CDC) recommends that everyone over the age of 6 months receive both doses of the mRNA COVID-19 vaccine (BNT162b2 and mRNA-1273)^1,2^. Large population-based studies have shown that a two-dose series of the mRNA COVID vaccine provides moderate protection against symptomatic infection in children^3^, as well as in the development of severe COVID-19-associated outcomes^4^. However, there is a lack of knowledge regarding how maternal vaccine timing, placental Fc-receptor binding and fetal characteristics drive transplacental antibody transfer and persistence of antibodies in infants over the first year of life.

While infants under the age of 6 months remain ineligible to receive the COVID-19 vaccine, they are one of the populations at highest risk for severe COVID-19 in the setting of SARS-CoV-2 infection. Infants younger than 6 months not only have higher COVID-19 hospitalization rates compared to other pediatric groups, but also are at higher risk for acquiring life-threatening complications from COVID-19, including acute respiratory failure ^5,6^. Maternal immunization is a key strategy to protect infants younger than 6 months from infectious diseases through passive transplacental antibody transfer ^7^. Additionally, maternal immunization shapes infant immune development by priming the cellular immune response ^8^. Although the level of antibody necessary to achieve infant protection against severe COVID-19 remains unknown, one study showed that the completion of the COVID-19 vaccination during pregnancy is correlated with reduced COVID-19 hospitalization rates in infants younger than 6 months of age, with greater protection for infants conferred when the mother received the COVID-19 mRNA vaccine after 20 weeks gestational age ^9–11^. More granular information on how timing of maternal vaccination influences infant antibody levels over the first year of life was previously not available.

Recent studies examining COVID-19 vaccination during pregnancy have demonstrated the transplacental antibody transfer of anti-Spike immunoglobin (Ig) G antibodies ^12–16^. While there are limited data on the persistence of maternal anti-Spike IgG antibodies in infants ^16,17^, no study has performed comprehensive profiling of maternal COVID-19 vaccine-induced antibodies that persist in infants through 12 months of age, including all IgG subclasses, Fc receptor (FcR) binding profiles, and against both SARS-CoV-2 wild-type and variants of concern (VOCs) including B.1.1.7 (Alpha), B.1.351 (Beta), B.1.617.2 (Delta), P.1 (Gamma), and B.1.1.529 (Omicron) variants. FcR binding is a critical consideration, as the Fc region of the antibodies plays an important role in protection against infection and the resolution of SARS-CoV-2 infection ^18,19^ by inducing the antiviral activity of the innate immune system ^20,21^. The Fc region of antibodies also plays a critical role in the transfer of antibodies across the placenta ^22,23^, and a more granular understanding of antibodies that persist in infants after maternal COVID vaccination may have broader implications for understanding optimal vaccine timing and features of vaccine-induced antibodies that maximize neonatal and infant protection. Such an understanding may be relevant to new vaccines with specific indications for pregnant individuals, including the RSV vaccine and others still in development. In this study, we performed unbiased systems serology to evaluate the levels and FcR binding profile of SARS-CoV-2-specific antibodies transferred from mother to infants, as well as their persistence in infants up to 12 months of age. Our analyses reveal differences in antibody profiles of infants whose mothers were vaccinated at different stages of gestation. This work further highlights the importance of understanding the timing of vaccination in pregnancy to provide optimal protection to susceptible infants during their first critical months of life.

## RESULTS

### Maternal COVID-19 vaccine-induced antibodies are detected in infants up to 12 months of age

To comprehensively profile COVID-19 vaccine-induced antibodies in infants up to 6 months of age, we evaluated 153 plasma/serum samples from a prospectively-collected cohort of 100 unique infants, including *N*=55 infants at two months, *N*=80 infants at 6 months, *N*=14 infants at 9 months, and *N*=4 infants at 12 months. Maternal *(N=*84) and cord (*N*=76) blood was available for a subset of the infants. During the course of the study, none of the included infants tested positive for COVID-19 (see Methods and Supplemental Figure 1). The mothers in this study all received both doses of the first generation (2020-2021) mRNA vaccinations (mRNA-1273 or BTN162b) or one dose of the Ad26.CoV2.S vaccine during pregnancy, and were not infected with SARS-CoV-2 prior to vaccination or during pregnancy. Table 1 details relevant clinical characteristics of mothers and infants in the study.

**Table 1:** Clinical characteristics of included infants. All included infants (N_p_ = 100) reflect the first maternal COVID-19 vaccination series. Some infants’ samples were collected at multiple time points, thus there are a total of 153 infant samples from 100 unique infants. All infants were born to mothers who were fully vaccinated in pregnancy as defined at the time (received both doses of the original COVID-19 mRNA vaccine series, or a single dose of the Ad26.COV2.S vaccine). None of the included infants had symptoms of COVID- 19 nor did they ever test positive for SARS-CoV-2.

|  | Overall<br>unique<br>infant<br>(N = 100) | 2 month-old infants<br>(N = 55) | 6 month-old infants<br>(N = 80) | 9 month-old infants<br>(N = 14) | 12 month-old infants<br>(N = 4) |
| --- | --- | --- | --- | --- | --- |
| **Vaccine Platform, N (%)** |  |  |  |  |  |
| mRNA-1274 | 31 (31) | 17 (31) | 28 (35) | 8 (57) | 1 (25) |
| BNT162b.2 | 59 (59) | 30 (55) | 48 (60) | 5 (36) | 3 (75) |
| Ad.26.COVS.2 | 10 (10) | 8 (15) | 4 (5) | 1 (7) | 0 (0) |
| **Infant Sex, N (%)** |  |  |  |  |  |
| Female | 49 (49) | 24 (44) | 41 (51) | 6 (43) | 3 (75) |
| **Gestational age at 1<sup>st</sup>**<br>**mRNA vaccine in**<br>**maternal series, N (%)** |  |  |  |  |  |
| < 20 weeks | 26 (29) | 23 (49) | 20 (26) | 1 (7) | 0 (0) |
| 20 – 28 weeks | 29 (32) | 16 (34) | 26 (34) | 7 (54) | 0 (0) |
| > 28 weeks | 35 (39) | 8 (17) | 30 (40) | 5 (36) | 4 (100) |
| **Gestational age at 2<sup>nd</sup>**<br>**mRNA vaccine in**<br>**maternal series, N (%)** |  |  |  |  |  |
| < 20 weeks | 15 (17) | 12 (25) | 13 (17) | 0 (0) | 0 (0) |
| 20 – 28 weeks | 27 (30) | 21 (45) | 22 (29) | 3 (23) | 0 (0) |
| > 28 weeks | 48 (53) | 14 (30) | 41 (54) | 10 (77) | 4 (100) |
| **Gestational age at**<br>**Ad26.COVS.2 Vaccine,**<br>**N (%)** |  |  |  |  |  |
| < 20 weeks | 3 (30) | 3 (37.5) | 0 (0) | 0 (0) | 0 (0) |
| 20 – 28 weeks | 2 (20) | 2 (25) | 1 (25) | 0 (0) | 0 (0) |
| > 28 weeks | 5 (50) | 3 (37.5) | 3 (75) | 1 (100) | 0 (0) |

We first plotted the Spike- and RBD-specific wild-type (WT) IgG antibody titer of mother-cord dyads, and 2-month-old and 6-month-old infants (Figure 1A). Expanding upon previous reports examining primarily total IgG against Spike ^14,24–26^, our results show cord samples are similar to maternal with respect to anti-Spike and anti-RBD IgG subclasses and IgG FcR binding responses. We observed that anti-Spike WT IgG1 was the most persistent antibody at 6 months of age, with 70% (56/80) of 6-month-old infants having detectable anti-Spike WT IgG1, while only 11% (9/80) had detectable IgG3 antibody levels, 2/80 with detectable anti-Spike IgG2, and no infants showing detectable levels of IgG4.

**Figure 1:**
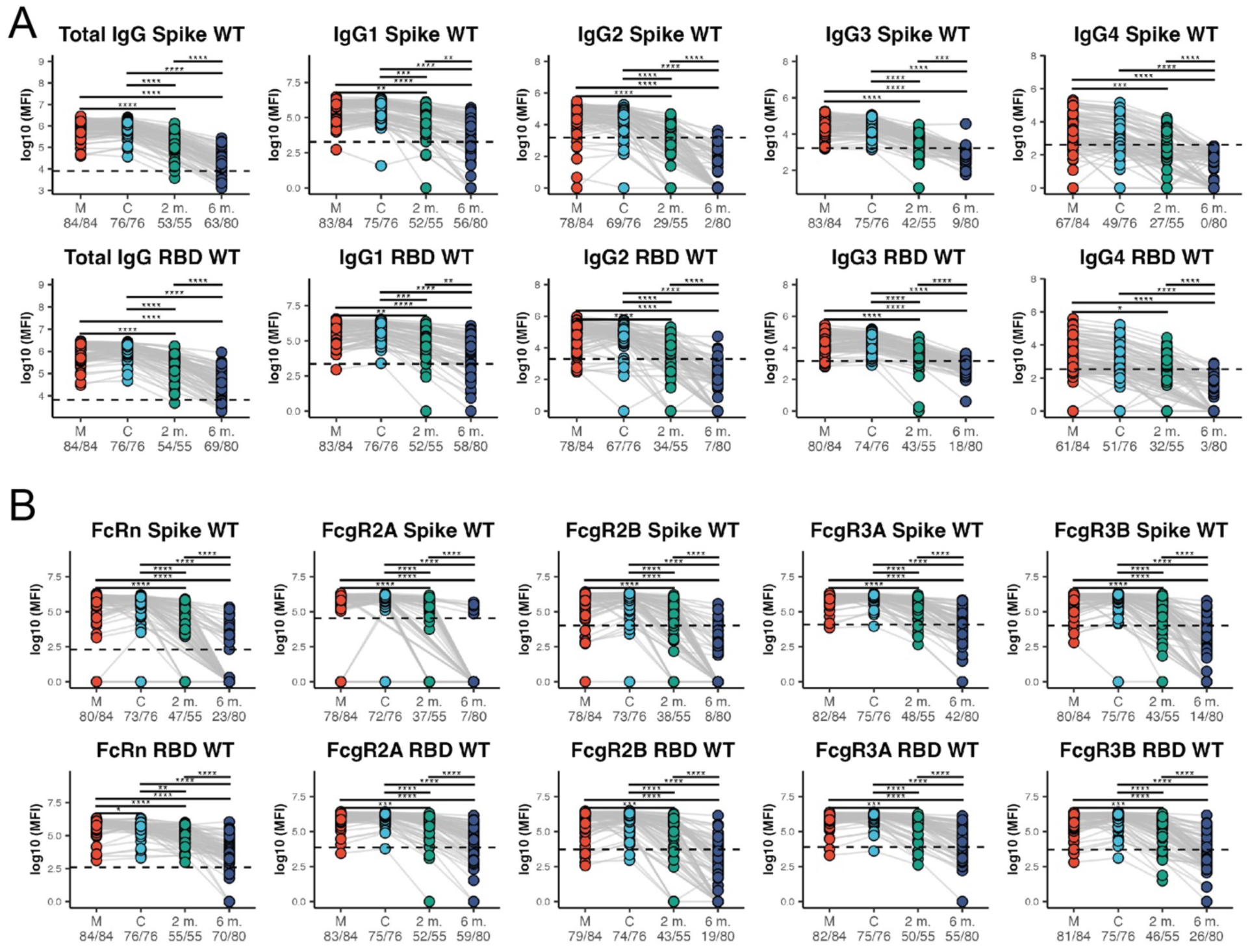
Maternal vaccine-induced SARS-CoV-2 antibodies persist in 2-month-old and 6-month-old infants. Dot plots showing **(A)** Spike-specific WT (top) and RBD-specific WT antibody levels and **(B)** FcR profiles of maternal (M), cord (C), 2-month-old infants (2 m), and 6-month-old infants (6 m). Significance was determined by a Kruskal-Wallis test followed by Benjamini-Hochberg procedure for multiple hypotheses correction. If statistically significant, a two-sided Mann-Whitney U test was performed. (*p<0.05, **p<0.01,***p<0.001,****p<0.0001). The dashed line represents the level of detection for each feature, defined as one standard deviation above the mean value for assay negative biological controls (SARS-CoV-2 negative, unvaccinated samples).

We also observed similar persistence trends between Spike- and RBD-specific antibodies. 73% (58/80) of 6-month-old infants had detectable anti-RBD WT IgG1, 23% (18/80) had detectable IgG3, 9% (7/80) had detectable IgG2, and 3/80 infants had detectable anti-RBD WT IgG4. These findings are overall consistent with previous reports of a transfer hierarchy of subclass-specific IgG transfer through the placenta from the mother to cord, with IgG1 > IgG3 > IgG4 > IgG2^27–29^.

Furthermore, we observed detectable Spike-specific and RBD-specific FcR-binding IgG levels in infants through 6 months of age (Figure 1B). Interestingly, 47/55 2-month infants demonstrated detectable antibody binding profiles against Spike-specific and 55/55 against RBD-specific neonatal Fc receptor (FcRn), with similarly high detection of antibodies binding canonical Fc-receptors FcγR2A and FcγR3B at 2 months of age (Figure 1B). However, greater differences in the persistence of the FcR-binding against Spike versus RBD in infants begin to emerge at 6 months of age. We observed only 29% (23/80) of infants had detectable antibodies binding Spike-specific FcRn, 9% (7/80) binding FcγR2A, 10% (8/80) binding FcγR2B, 53% (42/80) binding FcγR3A, and 18% (14/80) binding to FcγR3B. RBD-specific Fc-receptor binding was more persistent than that against Spike, with 88% (70/80) of infants having detectable RBD-specific antibodies binding FcRn, 74% (59/80) binding FcγR2A, 24% (19/80) binding FcγR2B, 69% (55/80) binding to FcγR3A, and 33% (26/80) binding to FcγR3B. Combined, these data demonstrate higher persistence of WT RBD-specific Fc-receptor binding IgGs compared to WT Spike at 6 months, with highest persistence of antibodies binding FcRn.

We also analyzed the levels of the same Spike- and RBD-specific WT antibodies in infants 9 months (*N*=12) and 12 months (*N*=4) of age (Supplemental Figure 2). We observed 10/12 infants at 9 months, and 2/4 infants at 12 months had detectable Spike-specific IgG1 levels. Similar persistence was observed for IgG1 against RBD (8/14 at 9 months, 1/4 at 12 months of age). There was negligible persistence of other IgG subclasses at 9 and 12 months of age. We also observed detectable FcR binding levels of RBD-specific antibodies only, with 13/14 9-month infants demonstrating detectable binding to FcRn, and 2/14 and 3/14 demonstrating detectable RBD-specific binding to FcγR3AV and FcγR2AR, respectively.

Of note, 12 infants whose mothers received the Ad26.CoV2.S vaccine were enrolled in the study and systems serology was performed on their samples, including 8 at 2 months, 3 at 6 months and 1 at 9 months. Of these 12, 75% (9/12) mothers were vaccinated at > 20 weeks’ gestation. These infants were included in all analyses except those pertaining to maternal timing of vaccine dose 1 and dose 2, which is only relevant to the mRNA vaccines (Supplemental Figures 3-5 Figures 3-4). Understanding antibody persistence in infants whose mothers received Ad26.CoV2.S may be relevant for future adenovirus-based vaccine engineering or viral-based therapeutics; we therefore report here that 6 / 9 infants had detectable IgG Spike-WT antibodies at 2 months, 1/3 infants at 6 months, and none at 9 months of age.

### Persistence of maternal COVID-19 vaccine-induced antibodies at 6 months of age differs by Variants of Concern

Next, we examined the cross-reactivity of the maternal COVID-19 vaccine induced antibodies and antibody FcR binding profile against wild-type SARS-CoV-2 Spike and RBD, as well as Spike and RBD VOCs in cord and infant samples (Figure 2A). Of note, the vaccines were administered to mothers between 12/18/20 and 6/22/21, and thus represent the first generation BNT162b2 and mRNA-1273 vaccines. To evaluate the persistence of the maternal vaccine-induced antibodies in infants against the VOCs, we compared the percent of infants in each group with detectable IgG subclasses and FcR binding profiles against Spike-specific and RBD-specific regions of the VOCs (Figure 2A).

**Figure 2:**
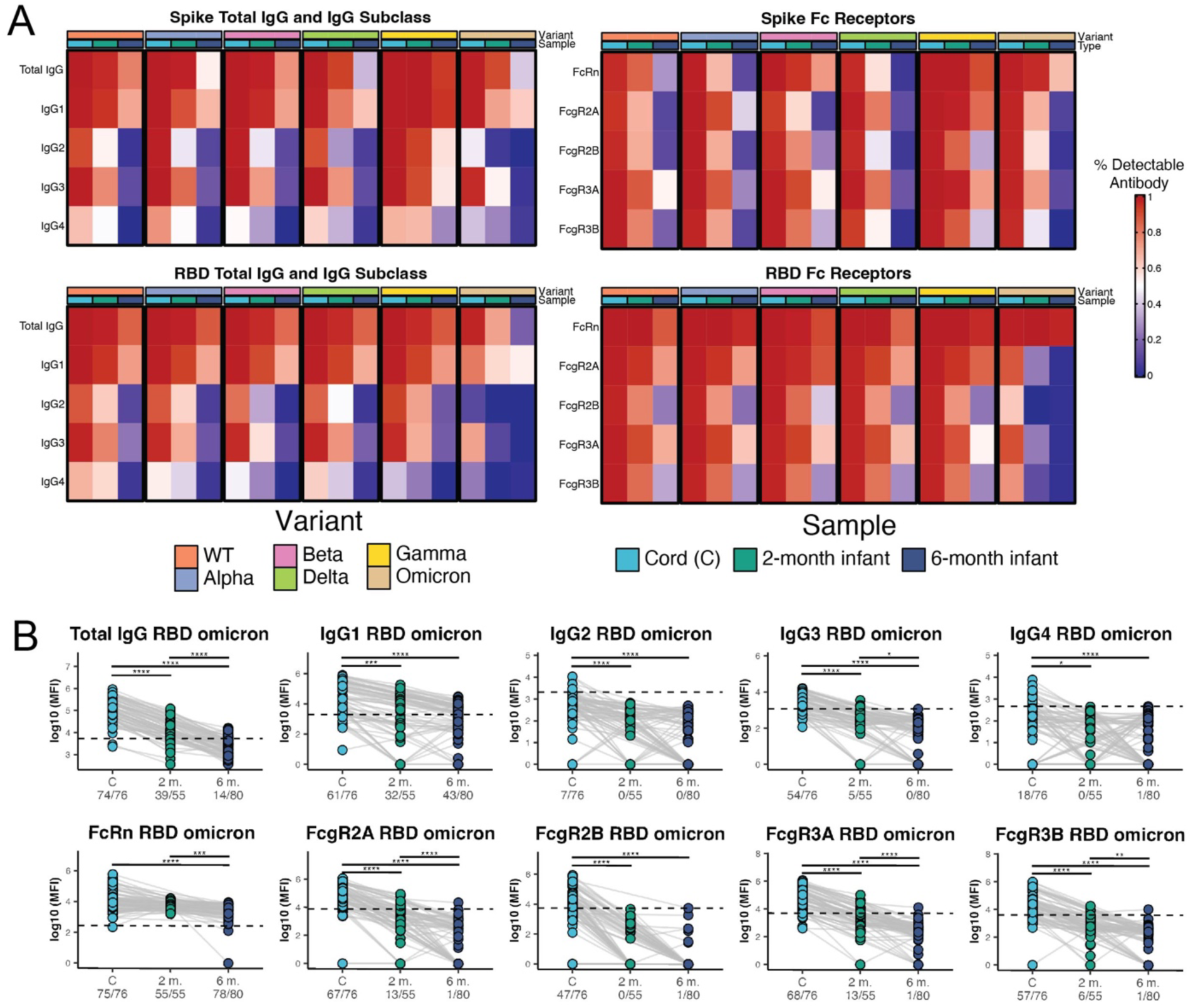
IgG subclasses and Fc-receptor binding capacity of antibodies against SARS-CoV-2 variants of concern in umbilical cord blood, 2-month-old, and 6-month-old infants (A) The heatmap shows the ratio of infants with detectable antibody titer or FcR binding levels N=76 for cord (C), N=55 for 2-month-old infants (2 M), and N=80 for 6-month-old infants (6 M) against the Spike or RBD region of wild-type (WT) strain and Variants of Concern (VOCs) including Alpha, Beta, Delta, Gamma, and Omicron. **(B)** Univariate plots showing RBD-specific Omicron antibody IgG levels (top) and FcR binding profiles in cord (C), 2-month-old infants (2 m), and 6-month-old infants (6 m) Significance was determined by a Kruskal-Wallis test followed by Benjamini-Hochberg procedure for multiple hypotheses correction. If statistically significant, a two-sided Mann-Whitney U test was performed. (*p<0.05, **p<0.01,***p<0.001,****p<0.0001). The dashed line represents the level of detection for each feature, defined as one standard deviation above the mean value for assay negative biological controls (SARS-CoV-2 negative, unvaccinated samples).

The first-generation COVID-19 mRNA vaccines generated antibodies cross-specific against most VOCs that persisted to 6 months, with highest persistence of antibodies cross-binding to Delta and to Gamma and lowest to Omicron. For all IgG subclasses and Fc receptor profiles apart from IgG2 and IgG4, we observed detectable levels of antibodies and FcR binding against either the RBD- or Spike-specific antigen of all VOCs except Omicron in infants up to 6 months of age (Figure 2A). These findings are consistent with previous work from our group, demonstrating that the Spike-specific vaccine-induced antibodies were able to robustly bind to other VOCS, with lower affinity observed for the RBD-specific region of the VOCs^30^. As seen for WT Spike and RBD (Figure 1A), IgG2 and IgG4 subclasses overall had low persistence, including against the different VOCs. While there was relatively high persistence of IgG1 at 6 months against Spike-specific Omicron (70% of infants showing detectable IgG1 antibodies), very few (N=9/80) 6-month infants had detectable IgG3 against Spike-specific Omicron, and only 14/80 6-month infants had detectable Spike-specific Omicron antibodies with Fc-receptor binding capabilities, with highest persistence of antibodies binding FcγR3A (8/80). While IgG1 against Omicron RBD persisted at 6 months in 54 % (43/80) of infants, a sharp decrease in the persistence of IgG3 against RBD-specific Omicron was observed, with only 5/55 infants demonstrating detectable levels of this antibody even at 2 months (Figure 2B). We also saw strikingly low persistence of RBD-specific Omicron FcR binding in 2- and 6-month infants, with only FcRn-binding antibodies persisting to any significant extent. The reduced ability of vaccine-generated antibodies to bind the highly-mutated^31^ RBD region of the Omicron Spike protein relative to other VOCs has been demonstrated previously^32^. The data presented here suggest that RBD-specific anti-Omicron antibodies also may engage non-canonical placental Fc-receptors to a lesser extent than antibodies against other VOCs, given their significantly reduced persistence in 2- and 6-month-old infants.

### The timing of maternal vaccination influences antibody levels in 2-month-old infants

Previous studies have demonstrated that timing of initiation of COVID-19 vaccination in pregnancy influenced the levels of maternal vaccine-induced antibodies at delivery and transplacental transfer to the umbilical cord^14,16,24,33,34^. Knowledge is lacking, however, regarding how the timing of not only the first, but also the second dose of the mRNA vaccine in pregnancy, impacts maternal antibody levels and persistence of antibody in infants. Thus, we wanted to examine specifically how the timing of both first and second mRNA vaccine dose impacted the profile of passively transferred maternal antibodies in infants. The 2-month cohort was selected for this subanalysis, given the higher and more uniformly persistent levels of all antibody features in this group, relative to the 6-month-old infant cohort.

First, we grouped the 2-month infant cohort into five groups, based on when the pregnant individual received dose 1 and dose 2 of the primary mRNA COVID-19 vaccination series (Figure 3A). For this analysis, we excluded infants whose mother received the Ad26.CoV.2 vaccine, as only a single dose of this vaccine was administered (*N*=8 2-month infants). Timepoints of < 20 weeks, 20-28 weeks and > 28 weeks were chosen as cut points for analysis, given previous work from the CDC demonstrating that maternal vaccination after 20 weeks’ gestation was more effective in preventing COVID-19-associated infant hospitalizations in the first 6 months of life, relative to when maternal vaccination was completed prior to 20 weeks^9,10^. The 20-28 week and >28 week window were selected to gain increased trimester-specific granularity on optimal timing of maternal vaccine administration.

**Figure 3:**
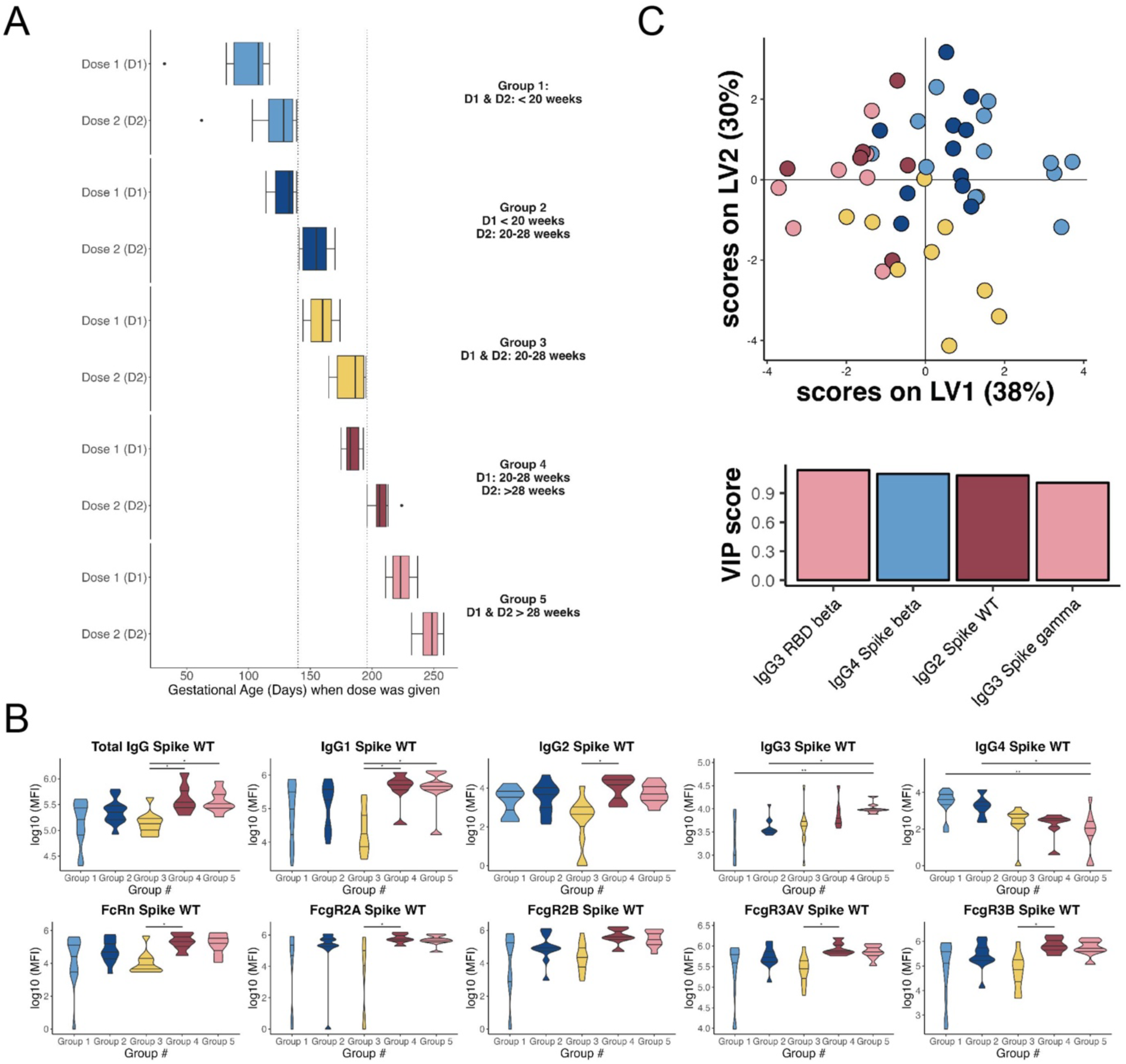
Maternal vaccine timing drives IgG levels and Fc-receptor binding profiles in 2-month-old infants. Infants whose mothers were vaccinated in the late second and early third trimester (received at least one dose of mRNA vaccine after 28 weeks) had the highest levels of antibodies at 2 months. **(A)** Diagram showing the grouping strategy for 2-month-old infants based on the Dose 1 (D1) and Dose 2 (D2) COVID-19 vaccination. Group 1: *N = 12*, Group 2: *N* = 11, Group 3: *N* = 10, Group 4: *N* = 6, Group 5: *N* = 8. **(B)** Univariate plots showing Spike-specific WT antibody and Fc receptor (FcR) levels of each 2-month-old infant group (as shown in in Figure 3A) Significance was determined by a Kruskal-Wallis test followed by Benjamini-Hochberg procedure for multiple hypotheses correction. If statistically significant, a two-sided Mann-Whitney U test was performed. (*p<0.05, ****p<0.0001). The middle line represents the median, whereas the other lines represent the first and third quartiles. **(C)** (Top) A partial-least squares discriminant model (PLSDA) was built using LASSO feature regularization and variable selection of the infant cohort group, resulting a reduced set of features that were consistently in at least 80 of the 100 LASSO models. (Bottom) Variable importance in projection (VIP) score of the selected features. The magnitude indicates the importance of the features in driving the separation in the model. The color of the feature corresponds to the group to which the feature was enriched.

Levels of Spike-specific IgG subclasses and Fc-receptor binding antibodies in 2-month-old infants by maternal vaccine timing are plotted in Figure 3B. These analyses demonstrated that for most subclasses and Fc receptor binding features, infants whose mother received at least one dose of mRNA vaccine after 28 weeks (Groups 4 and 5, maroon and rose groups) had the highest levels of maternal COVID-19 vaccine-induced antibodies. Interestingly, the lowest level of IgG1 and IgG2 subclasses and Fc receptor binding features was seen in infants whose mothers received both doses of the mRNA vaccine in the 20-28 week window. Given the prior data demonstrating enhanced protection against COVID-19-associated hospitalization when the vaccine series was completed after 20 weeks’ gestation^9,10^, we hypothesized we would observe the lowest antibody titers in infants whose mothers started and/or completed the entire vaccine series prior to 20 weeks. Instead, we observed that receiving both doses of the mRNA vaccine in the mid-to-late second trimester (Group 3, yellow) was associated with the lowest levels of anti-spike IgG1 and IgG2 in infants, which was consistent with prior work in a separate cohort, suggesting less robust maternal antibody responses in second trimester vaccination relative to first and third^14^. These less robust 20-28 week responses were hypothesized to be secondary to the known relative maternal immune quiescence of the second trimester, which occurs at least in part to facilitate rapid fetal growth^35^.

However, the finding that the lowest antibody levels were seen in infants whose mothers started and finished the vaccine in the 20-28 week window should be interpreted with an important caveat: Group 3 included 9 males and only 1 female infant, whereas other groups in this analysis were comprised by nearly equal numbers of males and females. Univariate analyses revealed that male infant sex was associated with significantly reduced levels of Spike-specific IgG2 antibodies at 2 months of age relative to female infants (Supplemental Figure 3A). We also observed reduced levels of Spike-Specific IgG1 antibodies in male infants compared to female infants, although this finding did not achieve statistical significance (Supplemental Figure 3A). Uniform Manifold Approximation and Projection Analysis (UMAP) also revealed antibody and Fc-receptor variation by timing of maternal vaccination and infant sex (Group 3, Supplemental Figure 3B). It is therefore possible that the lower titers noted in infants after late second trimester maternal vaccination are a function of either maternal vaccine timing, male infant sex, or both.

Finally, we observed that infants whose mother received at least one dose of mRNA vaccine prior to 20 weeks’ gestation had the highest levels of IgG4. Previous work demonstrated that IgG4 antibodies after COVID-19 vaccination were less inflammatory and had more muted effector functions (antibody-dependent phagocytosis and complement deposition) than other subclasses, and IgG4 levels rose with longer interval from 2^nd^ mRNA vaccination^36^. Thus, the elevated IgG4 levels in infants whose mothers were vaccinated at < 20 weeks may be present due to increased passage of time and increased opportunity for antibody class switching in the mother, and these antibodies may have more muted effector functions and therefore may be less desirable to pass to infants than other subclasses of IgG. Consistent with this concept, we observed the highest levels of IgG3, the subclass most associated with an effective early anti-viral response^37^ in infants whose mothers received the first vaccine dose after 20 weeks’ gestation.

Given the observed differences in infant antibody levels by timing of maternal vaccination, we next sought out to determine the combination of features that best separated infants based on timing of vaccination using partial least squares discriminant analysis (PLSDA), followed by least absolute shrinkage and selection operator (LASSO) feature regularization to prevent overfitting (Figure 3C). The LASSO-selected features that are important (based on Variable Importance in Projection (VIP) analyses) in building a model to distinguish groups based on maternal vaccine timing are highlighted in Figure 3C, and include not only the Spike-specific WT antibody levels and FcR binding profiles, but also some RBD-specific and VOC-specific features. Other features used in building the PLSDA model, as well as the model validation results, are depicted in Supplemental Figure 4A and 4B. Among the top features that distinguish the groups, three (anti-RBD Beta IgG3, anti-Spike gamma IgG3, and anti-Spike WT IgG2) were enriched in either Group 4 or Group 5 (at least one vaccine dose administered > 28 weeks), and 2 features (anti-Spike beta IgG4 and anti-RBD delta IgG4) were enriched in Group 1 or 2 (at least one vaccine dose administered < 20 weeks, Figure 3C and Supplemental Figure 4A). Similar to levels of IgG subclasses, numerous FcR binding features were enriched in Group 4 or Groups 4 and 5, primarily relative to Group 3 (fully vaccinated 20-28 week window).

Given that LASSO selects a minimal set of features to prevent overfitting of the data, we generated a correlation heatmap to determine additional features that correlate with the top features used in the PLSDA model (Supplemental Figure 4C). IgG3 and IgG4 features, regardless of antigen target, were correlated with other IgG3 and IgG4 antibodies, respectively (Spearman correlation rho > 0.75). Spike- and RBD-specific IgG3 and Spike-specific IgG2 both were positively correlated with FcR binding levels. Overall, these analyses demonstrated that the timing of not only the 1^st^ but also the 2^nd^ dose of maternal COVID-19 vaccination influence antibody subclass levels in 2-month-old infants.

To evaluate the impact of maternal vaccine timing on antibody features in 6-month infants, we sought to determine a set of features that best separate 6-month infants based on maternal vaccination timing, as was done with the 2-month infant cohort, using LASSO feature regularization in conjunction with PLSDA classification (Supplemental Figures 5A-C). Similar to the results in 2-month infants, the LASSO-selected features show an enrichment of antibody levels in 6-month infants whose mother was vaccinated > 28 weeks (Supplemental Figures 5B and 5C). Although the univariate plots of these features did not demonstrate statistically significant differences, these plots demonstrate that infants whose mothers received the vaccine > 28 weeks had elevated antibody and Fc-receptor binding levels compared to those vaccinated <20 or 20-28 weeks (Supplemental Figure 5D).

### Evaluating the persistence of maternal-vaccine-induced antibodies in 6-month infants by timing of the maternal COVID-19 vaccine series in early versus later 3^rd^ trimester: implications for RSV vaccination in pregnancy

Recently, the CDC’s Advisory Committee on Immunization Practices recommended the bivalent recombinant respiratory syncytial virus prefusion F protein (RSV preF) vaccine for seasonal use in pregnant individuals at 32-36 weeks’ gestation, to prevent RSV lower respiratory tract disease in infants^38^. In a large randomized controlled trial of over 7300 pregnant individuals, RSV vaccination from 24-36 weeks was associated with protection efficacy of 69.4% against severe lower respiratory tract disease in infants at 6 months of age^39^. However, RSVpreF vaccination was ultimately approved in pregnant individuals in the narrower window of 32-36 weeks’ gestation. As this narrower and later window was not explicitly studied in the trial, and given our findings that timing of maternal vaccination administration had significant impact on antibody persistence in infants, we sought to examine antibody persistence in infants at 6 months with maternal administration of the COVID-19 vaccines at 28-32 weeks versus 32-36 weeks’ gestation in our sample. While this is a different vaccine platform with a two-dose rather than one dose regimen, given the lack of data on antibody persistence in infants when maternal vaccines are administered at 32+ weeks of gestation, these data can add significantly to existing knowledge.

First we evaluated antibody features in 6-month-old infants as a function of timing of maternal vaccination in pregnancy using polar plots (Figure 4A). Because of their low persistence in the 6-month infant cohort (Figure 1A), we excluded IgG2 and IgG4 from these analyses (overall more than 70% of the samples for IgG2 and IgG4 fell below the limit of detection threshold). Comparing the mean percentile ranks of antibody subclasses and Fc-receptor binding features persistent in 6-month-old infants by maternal vaccine timing in pregnancy, it was clear that maternal vaccination at > 28 weeks was associated with the most robust persistence of antibodies in 6-month-old infants (relative to < 20 weeks or 20-28 weeks). These results were thus consistent with the prior results for 2-month-old infants (Figure 3).

**Figure 4:**
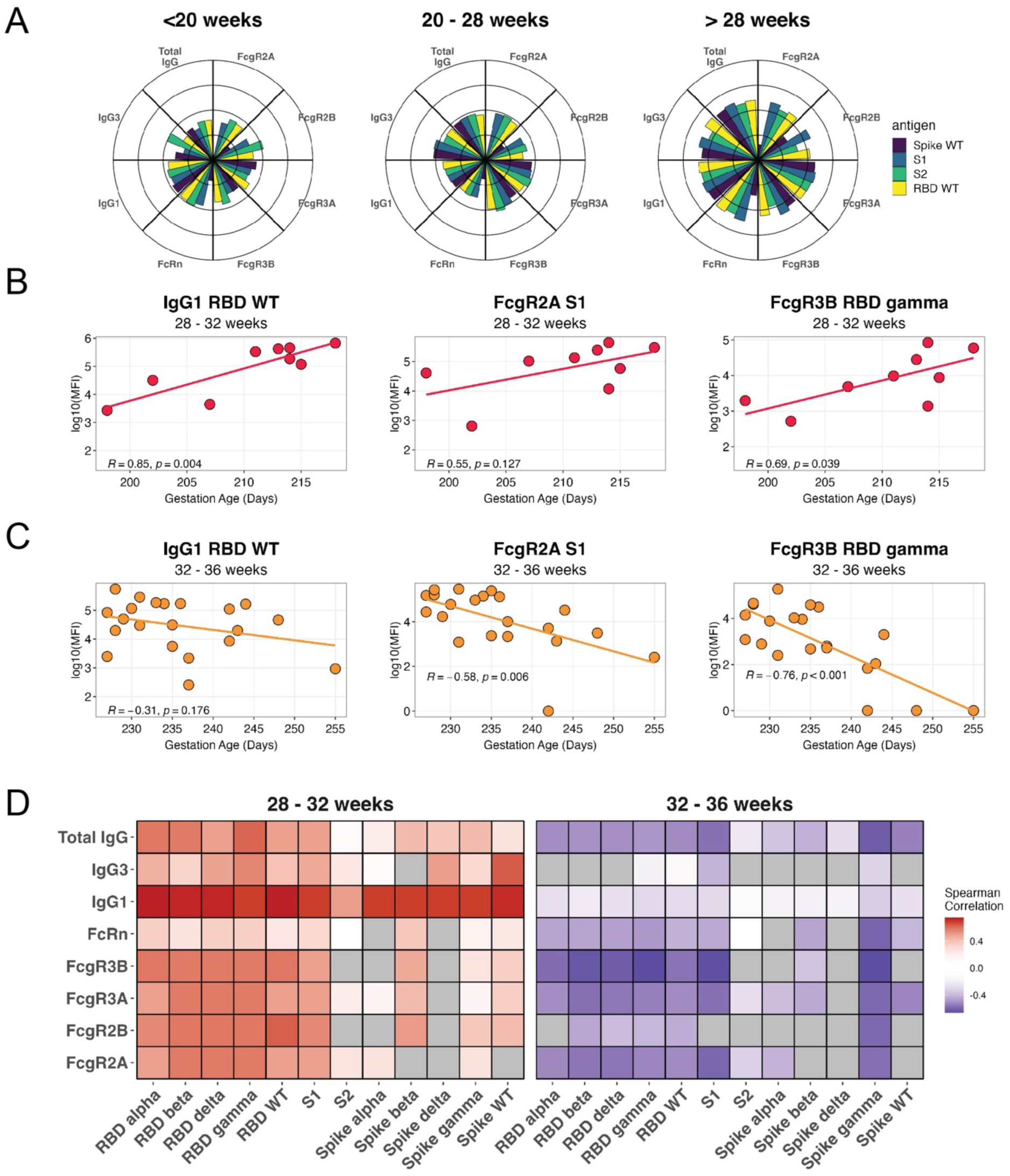
Maternal vaccine administration at > 32 weeks is associated with reduced antibody persistence in 6-month-old infants. **(A)** The polar plots depict the mean percentile ranks of the antibody features within 6 month infants whose moms were vaccinated with the first dose of the mRNA COVID-19 vaccine <20 weeks (*N*=20), 20 – 28 weeks (*N*=26), and > 28 weeks (*N*=30). Each wedge represents an antibody feature, and the size of the wedge depicts the mean percentile ranging from 0-1. **(B)** Linear regression models fits the relationship between gestation age of dose 1 (days) (those vaccinated between 28 – 32 weeks) and anti-RBD WT IgG1, anti-S1 FcγR2AR, and anti-RBD gamma FcγR3B. The rho and p-values are reported in the plot **(C)** Linear regression models fits the relationship between gestation age of dose 1 (days) (those vaccinated between 32 - 36 weeks) and anti-RBD WT IgG1, anti-S1 FcγR2AR, and anti-RBD gamma FcγR3B. The rho and p-values are reported in the plot. **(D)** The heatmap shows the Spearman correlation score for each feature against gestational age (days) within infant samples whose mothers received the first dose of the mRNA COVID-19 vaccine between 28 – 32 weeks or 32 – 36 weeks. The red boxes indicate samples with a positive Spearman correlation, blue boxes indicate samples with a negative Spearman correlation. Within each group, gray boxes are features which were excluded from the analysis (if >70% of values fell below the threshold of detection, defined as less than one standard deviation above the mean of SARS-CoV-2 negative, unvaccinated control samples).

To parse more specifically how timing of maternal vaccination **within** the 3^rd^ trimester influenced persistence of infant antibody titers at 6 months, we then evaluated levels of IgG subclasses and Fc-receptor binding antibody levels in 6-month infants as a function of maternal vaccination from 28-32 weeks (Figure 4B), versus maternal vaccination from 32-36 weeks (Figure 4C). We found that the majority of antibody features, including IgG subclasses and Fc-receptor binding, were significantly positively correlated with gestational age at maternal vaccine administration across the 28-32 weeks’ gestation period, particularly IgG1 against RBD and Spike WT and VOCs, as well as FcγR2AR-, FcγR3AV- and FcγR3B-binding antibodies (Figures 4B, 4D). In contrast, many of the same features decreased steadily as a function of time across the vaccine window when maternal vaccination was initiated from 32-36 weeks (Figures 4C, 4D). In summary, these analyses demonstrate a significant positive correlation between infant antibody levels at 6 months and gestational age at vaccination across the 28-32 week gestational age window. However, this relationship reverses across the 32–36 week window, with a significant negative correlation between 6-month infant antibody levels and gestational age at maternal vaccination These limited data suggest that the efficiency of transplacental transfer of maternal antibodies and durability in infants through 6 months of age may be reduced across the 32-36 week gestational age window.

## DISCUSSION

Maternal vaccination against COVID-19 has been shown to effectively reduce COVID-19 related hospitalization rates in infants younger than 6 months ^9,10^, through the transplacental transfer of maternal COVID-19 vaccine induced antibodies ^16,17^. Here, using systems serology, we profiled maternal antibodies in 100 infants up to 12 months of age in unprecedented detail. These experiments quantified not only total IgG level, but IgG subclass and FcR binding profile of antibodies against wild type Spike and RBD epitopes of SARS-CoV-2, as well as a wide range of variants of concern including Omicron. Although the COVID-19 pandemic is ongoing and protecting both mothers and vulnerable infants under 6 months of age who cannot yet be vaccinated against COVID-19 remains critical, our findings have relevance far beyond COVID-19. Our findings that antigen specificity (e.g., Spike versus RBD), IgG subclass (e.g. IgG1 and IgG3), and canonical (FcRn) versus non-canonical (Fcγ) placental receptor binding work in concert to drive persistence of antibodies in infants through 6 months of age may have implications for both future vaccine development and timing of administration in pregnancy. Our findings that both timing of maternal vaccination and fetal sex were significantly associated with persistence of maternally-transferred antibody in infants may be used to guide both pregnancy vaccination strategies, and our understanding of infant protection. Finally, our granular examination of the impact of maternal vaccine timing within the mid to late 3^rd^ trimester demonstrates enhanced antibody persistence in 6-month infants when vaccine occurs across the 28-32 week window, yet reduced persistence across the 32-36 week window, with potential immediate relevance for maternal RSV vaccination strategies. Thus, the detailed antibody map presented here provides the basis for a more mechanistic understanding of maternal antibody generation, transplacental transfer, and persistence of antibodies across the mother-infant dyad.

While limited data are available regarding the persistence of maternal COVID-19 vaccine-induced antibodies in infants^16,17^ this represents the largest study examining infant antibody persistence, with comprehensive infant follow up at 2, 6, 9 and even 12 months of age, albeit with more limited numbers profiled after 6 months. In addition, these represent the only available data regarding the persistence of IgG subclasses and FcR binding profiles in infants through 12 months of age. Our observation of IgG1 and IgG3 having the highest persistence in 6 month infants is consistent with the known transfer hierarchy of subclass-specific IgG transfer through the placenta from the mother to cord, with IgG1 > IgG3 > IgG4 > IgG2^27–29^. Subclass-specific IgG transfer hierarchy is mediated not only by the relative abundance of each subclass, but also FcR expression levels and Fc-FcR interaction affinity^28^. As an example, IgG3, one of the most persistent antibodies in 6- month infants, has high binding affinity to placental FcγR3A^40^. In addition to facilitating IgG3 transfer across the placenta, the finding that the most persistent FcR binding profiles at 6 months were for antibodies binding FcγR3A suggests preferential transfer of natural killer (NK) cell-activating antibodies^23,41^ to arm neonates with the most effective immunity from birth, as NK cells play a crucial role in neonatal first-line innate immune defenses^42^.

With respect to timing of maternal vaccination in pregnancy, prior epidemiologic studies demonstrated that maternal COVID-19 vaccination after 20 weeks’ gestation conferred the greatest infant protection against COVID-19-associated hospitalizations in the first 6 months of life^9,10^. These studies did not examine antibody levels in infants, nor did they examine the impact of vaccination timing within the second half of pregnancy on infant protection. While there are limited data available on antibody levels in infants after maternal COVID-19 vaccination^16,17^, these studies have not examined how timing of vaccination across a range of trimesters or within trimester impacts infant antibody levels. There are also no data on relative persistence of different IgG subclasses, nor on FcR binding, or activity against VOCs in infants within the first year of life. Our finding of the highest levels of IgG1-3 and FcR binding in infants whose mother was vaccinated with at least one dose of the mRNA vaccine after 28 weeks of gestation, and significantly lower IgG levels and FcR binding in infants born to mothers fully vaccinated in the 20-28 week window, builds upon our prior work demonstrating enhanced IgG levels, FcR binding and functions when mothers are vaccinated in 3^rd^ relative to 2^nd^ trimester^14,43^. The finding that IgG4 was highest in infants born to mothers vaccinated prior to 20 weeks likely can be at least partially explained by antibody class switching and a time effect, with < 20 week vaccinees having more time for class-switching to IgG4 and transfer of Ig4 across the placenta. IgG4 may be a less advantageous antibody subclass for newborns and early infants with immature immune systems, given that IgG4 responses are T-cell dependent^14,43^ and have reduced effector functions (e.g., antibody-dependent cellular phagocytosis and complement deposition)^44,45^.

Also relevant to considerations of timing of maternal vaccination is our focused subanalysis examining durability of placentally-transferred antibodies in 6-month-old infants after maternal vaccination in mid- to late-third trimester. This question is timely given the recent recommendation by the FDA and CDC/ACIP to restrict maternal RSV vaccination to the 32-36 week window of pregnancy^46–48^. Our data suggest that most effective transfer occurs in the context of maternal vaccination in the early third trimester (28-32 weeks), with progressively less efficacious placental transfer (reflected in lower levels of IgG and Fc-receptor binding) as vaccination occurs from 32-36 weeks. These limited data point to maternal RSV vaccination in the earlier part of the 32-36 week window potentially being more optimal to maximize infant protection through 6 months of age. These data should be interpreted with caution as the RSV vaccine is a different platform (recombinant protein) and a one-rather than two-shot regimen, with likely different transfer kinetics. However, given the current dearth of evidence to guide vaccine timing within the recommended window, we offer these analyses while we await further prospective data from pregnant RSV vaccine recipients and their infants.

With respect to the impact of infant sex on infant antibody persistence at 2 and 6 months of age after maternal vaccination, our finding that male infants had reduced persistence of antibodies at 2 and 6 months relative to females resonates with prior published data. Prior work by our group found that pregnant individuals carrying male infants had lower maternal levels of COVID-specific antibodies and reduced FcR binding antibody levels, compared to individuals carrying female infants after natural COVID infection^49^. Sex-specific anti-COVID antibody glycan profiles and sex-specific patterns of placental Fc-receptor expression that may be less favorable for transplacental antibody transfer to male infants were also previously reported^49^. Maternal immune response, specific antibody properties such as glycan profiles, and placental FcR expression all are influenced by fetal sex, making this a critical area for further research.

Lastly, we examined whether maternal vaccination with the first-generation COVID-19 vaccines induced antibodies against other VOCs, persistent in 2 and 6 month old infants, finding the most consistent cross-immunity was induced for the gamma and delta variants, but with persistent levels of antibodies and Fc receptor engagement against all VOCs in infants up to 6 months of age with the exception of Omicron. Persistence of Omicron RBD-specific IgG3 was negligible, with nearly no detectable levels of this antibody in even 2-month infants. Similarly, there was strikingly low Omicron RBD-specific Fcγ-receptor binding in 2- and 6-month infants in comparison to other VOCs or even to Omicron Spike-specific Fcγ-receptor binding. This could be due to the disproportionately high mutation burden in the RBD region of the Omicron Spike protein^31^. The extent to which these potential deficits in activity against Omicron could be overcome by infant or childhood vaccination with updated Omicron-specific COVID-19 vaccines is an important question. Additionally, it is likely that maternal vaccination with an updated COVID-19 booster against Omicron could augment transfer of Omicron Spike- and RBD-specific antibodies to infants, and this is a critical area of ongoing investigation^50^. Finally, these data highlight the importance of comprehensive antibody profiling to better understand the complexity of immunity transferred to the infant by maternal vaccination; such data clearly have implications for vaccine design and/or implementation strategies, particularly against pathogens with high mutation rates.

These results demonstrate that COVID-19 vaccination during pregnancy provides not only a robust antibody response in the mother, but antibodies detectable in infants through > 6 months of age. A fine-grained understanding of antibody subclasses and Fc-receptor binding profiles that persist in infants through 1 year of life is critical not only for understanding protection afforded infants by maternal COVID-19 vaccination, but for the development of rational vaccines and data-driven maternal vaccination strategies. Our findings related to the critical impact of vaccine timing across gestation and of infant sex on antibody durability at 2 and 6 months of age demonstrate the importance of ongoing investigations of these unique aspects of maternal-fetal immunity that should inform vaccine development, and can drive strategies to optimize maternal and neonatal immunity against a wide array of pathogens.

## Data Availability

All data in the present study are available upon reasonable request to the authors.

## ACKNOWLEDGEMENTS

Funding sources: Merck Sharp & Dohme Corp Investigator-Initiated Studies grant (MISP 61299, to A.G.E.); NIH/NIAID 1U19AI167899-01 to D.A.L., A.G.E., M.A.E., B.D.J.; NIH/NHLBI 5K08HL143183 to L.M.Y.

## AUTHOR CONTRIBUTIONS

A.G.E., P.A.L., B.D.J., N.N., D.A.L developed the concept, designed the experiments, and analyzed and interpreted the data. A.G.E., D.A.L., M.A.E., and B.D.J. obtained funding. N.N, and T.C. performed experiments. P.A.L and D.A.L performed the statistical modeling. P.A.L. and A.G.E. wrote the main paper. S.B., R.M.D., S.D., M.K.K., and C.D.P. collected and prepared samples. L.L.S., M.D.B., O.J., B.A., L.M.Y., K.J.G., A.G.E. recruited and enrolled participants. O.J., M.D.B., B.A., K.J.G., L.M.Y., and A.G.E. assisted with clinical data collection. M.A.E. gave conceptual advice and edited the manuscript. All authors approved of the final manuscript.

## STAR Methods

### RESOURCE AVAILABILITY

#### Lead Contact

Requests for information should be directed to the lead contact, Andrea G. Edlow

#### Material Availability

This study did not generate new unique reagents.

#### Data and Code Availability

The dataset generated during this study is available upon reasonable request. Code used for the analyses can be found at Mendeley Data: http://dx.doi.org/10.17632/35zy6p92f6.1

### EXPERIMENTAL MODEL AND STUDY PARTICIPANT DETAILS

#### Study Population

The study included *N*=108 unique infants born to mothers who received their initial COVID-19 vaccine series in pregnancy (dates of maternal vaccination Dec 18, 2020-June 22, 2021). Mothers enrolled their infants in this study conducted from July 23, 2021 to July 1, 2022. Ultimately samples collected from pregnant individuals infected with SARS-CoV-2 before vaccination or babies known to be infected with SARS-CoV-2 prior to sample collection were excluded (Supplemental Figure 1). Matched maternal and umbilical cord serum samples were collected at birth for a subset of mothers co-enrolled in a complementary pregnancy cohort (MGB IRB # 2020P003538). Infant capillary serum samples were collected via microneedle device (TAP^®^ II blood collection device, YourBio Health, Medford, MA) at 2 months, 6 months, 9 months and 12 months after birth for infants of vaccinated mothers. All infants were singletons except for 2 pairs of twins. All included pregnant participants signed informed consent for sample collection, and birthing parents signed consent for their infants (MGB IRB #s 2020P003538 and 2020P000955).

### METHOD DETAILS

#### Antigens

Spike and RBD proteins of SARS-CoV-2 D614G or variants of concern, along with S1 and S2were obtained from Sino Biological. These antigens were expressed in mammalian HEK293 cells. To biotinylate these antigens, NHS-Sulfo-LC-LC kits were used following the manufacturer’s instructions (ThermoFisher Scientific).

#### Multiplexed Luminex assay

Antigen-specific antibody levels and Fcy receptor binding were measured using a custom multiplexed Luminex assay, as previously described^51^. To couple the antigens to magnetic carboxylated fluorescent Luminex beads (Luminex Corporation), carbodiimide-NHS ester coupling was performed, with one bead region assigned to each antigen. The beads were first activated in an activation buffer, which consisted of 100 mM monobasic sodium phosphate (pH 6.2), 50 mg/ml N-hydroxysulfosuccinimide (Sulfo-NHS; Pierce) resuspended in H_2_0, and 1-ethyl- 3-[3-dimethlyaminopropyl]carbodiimide-HCl (EDC; Pierce) resuspended in the activation buffer. Following a 30-minute incubation at room temperature (RT), the activated beads were washed in coupling buffer (50 mM morpholineethanesulfonic acid, MES; pH 5.0) and then incubated with antigens for 2 hours at RT. Subsequently, the beads were blocked during a 30-minute incubation at RT in phosphate-buffered saline (PBS)-TBN (containing 0.1% bovine serum albumin [BSA], 0.02% Tween 20, and 0.05% azide [pH 7.4]). Lastly, PBS-Tween (0.05% Tween 20) was used to wash the beads coupled with the proteins, then the beads were stored in PBS with 0.05% sodium azide at 4°C.

#### Antigen-specific immunoglobulin quantification

Antigen-coupled beads were incubated with different sera dilutions (between 1:50 and 1:100, depending on the secondary reagent) at 4°C for 18 hours of shaking. After the incubation, the beads were washed three times with PBS-Tween (0.05% Tween 20) and mixed to phycoerythrin (PE)-conjugated mouse anti-human antibodies (IgG, IgG1, IgG2, IgG3, IgG4, IgA1, IgA2, or IgM) from Southern Biotech at a concentration of 1.3 μg/ml. Following an additional 1-hour incubation at RT with shaking, the beads underwent another three washes with PBS-Tween (0.05% Tween 20) and were then resuspended in sheath fluid (Luminex Corporation). To ensure accuracy, each serum sample was tested in duplicate. The levels of PE median fluorescence intensity (MFI) were measured using the iQue screener plus (Intellicyt) analyzer.

#### Antigen-specific Fcy-receptor binding

To investigate the relative binding levels of antibodies to individual Fcγ-receptors (FcγRs), Avi- tagged FcγR2AR (used to measure FcγR2A), FcyR2B (used to measure FcγR2B), FcγR3AV (used to measure FcγR3B), and FcyR3B were obtained from Duke Human Vaccine Institute^51^ . Biotinylation of these receptors was carried out using a BirA biotin-protein ligase (BirA500; Avidity), and any excess biotin was removed using Zeba spin desalting columns (7K MWCO; Thermo Fisher Scientific). Serum samples were diluted at a ratio of 1:500 and mixed with antigen-coupled beads to form immune complexes. Following an 18-hour incubation at 4°C, the beads were washed three times with PBS-Tween (0.05% Tween 20). Streptavidin-R-phycoerythrin (ProZyme) was then added to each biotinylated FcyR at a 4:1 molar ratio and allowed to incubate for 20 minutes at RT. Fluorescent labeled FcyRs (at a concentration of 1 μg/ml in 0.1% BSA-PBS) were subsequently added to the immune complexes, followed by an incubation of 1 hour at RT. Lastly, three washes with PBS-Tween (0.05% Tween 20) were done, then beads were resuspended in sheath fluid (Luminex Corporation) for analysis. The median PE intensity was measured using the iQue screener plus (Intellicyt) system, and all samples were tested in duplicate.

### QUANTIFICATION AND STATISTICAL ANALYSIS

Analyses were divided between mothers vaccinated at < 20 weeks’ gestation, versus 20-28 weeks (window fully in the 2^nd^ trimester) versus 28 weeks to delivery (3^rd^ trimester) to reflect the known increased protection against severe infant COVID-19 disease conferred by maternal vaccination after 20 weeks’ gestation (e.g. < 20 weeks, > 20 weeks)^9,10^, and to further subset vaccination after 20 weeks’ gestation into biological windows that coincide with the trimesters of pregnancy. All statistical analyses and figure generation were performed using R (version 4.1.0). For Luminex samples, all features were subtracted by phosphate-buffered saline (PBS) control values and were log_10_-transformed. As in previous studies^17^, detectable Luminex-derived features were defined as any value greater than the sum of the mean of the SARS-CoV-2 unvaccinated negative control adult samples (collected before the pandemic) and one standard deviation of those samples.

For univariate analyses of differences between maternal, cord, and infant samples, and as well as 2-month-old infant groups that vary between vaccination timing, significance was determined by Kruskal-Wallis followed by posthoc Benjamini-Hochberg adjustment. For analysis of the transfer between maternal and cord dyads, significance was calculated by a Wilcoxon signed-rank test followed by Benjamini-Hochberg. Multivariate analyses of the systems serology data were performed in R (version 4.2.1). All log10 data were then centered and scaled. Luminex-derived features for which 70% of values fell below one standard deviation above the mean of SARS-CoV-2 unvaccinated negative control samples were pruned. Uniform Manifold Approximation and Projection (UMAP), unsupervised dimensionality reduction technique^52^, was performed to evaluate the effect of maternal vaccination timing and fetal sex. Prior to UMAP analysis, the first 20 principal components (PCs) that explain 95% of the data were determined using principal component analysis (PCA)^53^. These PCs were then mapped used to reduce the high-dimensional serological data into a two-dimensional space with fine tuned parameters (n.neighbor = 30, min.dist =0.2). UMAP was performed using the R package ‘Seurat’.

For Partial Least Squares Discriminant Analysis (PLSDA) classification models, Least absolute shrinkage and selection operator (LASSO) feature selection^54^ was performed to extract the most significant features in distinguishing our groups. LASSO feature selection was implemented in the function select_lasso in the systemseRology package (version 1.0) and was performed 100 times, and only features that were chosen repeatedly were further analyzed in downstream analyses. PLSDA was performed using the LASSO-selected features. To evaluate model robustness, the permuted control models were first built by randomly shuffling the labels 100 times. These control models were then cross-validated 10 times to determine the accuracy of our model. Robustness was then calculated as the effect size of the actual and control distributions and defined as the exact p-values of the tail probabilities of the actual distributions within the control distributions.

## REFERENCES

1. CDC (2022). CDC recommends COVID-19 vaccines for young children (US Department of Health and Human Services, CDC).

2. CDC (2023). Interim Clinical Considerations for Use of COVID-19 Vaccines in the United States (US Department of Health and Human Services, CDC).

3. Powell, A.A., Kirsebom, F., Stowe, J., McOwat, K., Saliba, V., Ramsay, M.E., Lopez-Bernal, J., Andrews, N., and Ladhani, S.N. (2022). Effectiveness of BNT162b2 against COVID-19 in adolescents. Lancet Infect. Dis. 22, 581–583. 10.1016/S1473-3099(22)00177-3.

4. Klein, N.P., Stockwell, M.S., Demarco, M., Gaglani, M., Kharbanda, A.B., Irving, S.A., Rao, S., Grannis, S.J., Dascomb, K., Murthy, K., et al. (2022). Effectiveness of COVID-19 Pfizer-BioNTech BNT162b2 mRNA Vaccination in Preventing COVID-19–Associated Emergency Department and Urgent Care Encounters and Hospitalizations Among Nonimmunocompromised Children and Adolescents Aged 5–17 Years — VISION Network, 10 States, April 2021–January 2022. MMWR Morb. Mortal. Wkly. Rep. 71, 352–358. 10.15585/mmwr.mm7109e3.

5. Hamid, S., Woodworth, K., Pham, H., Milucky, J., Chai, S.J., Kawasaki, B., Yousey-Hindes, K., Anderson, E.J., Henderson, J., Lynfield, R., et al. (2022). COVID-19–Associated Hospitalizations Among U.S. Infants Aged <6 Months — COVID-NET, 13 States, June 2021–August 2022. MMWR Morb. Mortal. Wkly. Rep. 71, 1442–1448. 10.15585/mmwr.mm7145a3.

6. Hobbs, C.V., Woodworth, K., Young, C.C., Jackson, A.M., Newhams, M.M., Dapul, H., Maamari, M., Hall, M.W., Maddux, A.B., Singh, A.R., et al. (2022). Frequency, Characteristics and Complications of COVID-19 in Hospitalized Infants. Pediatr. Infect. Dis. J. 41, e81–e86. 10.1097/INF.0000000000003435.

7. Marchant, A., Sadarangani, M., Garand, M., Dauby, N., Verhasselt, V., Pereira, L., Bjornson, G., Jones, C.E., Halperin, S.A., Edwards, K.M., et al. (2017). Maternal immunisation: collaborating with mother nature. Lancet Infect. Dis. 17, e197–e208. 10.1016/S1473-3099(17)30229-3.

8. Marchant, A., and Kollmann, T.R. (2015). Understanding the Ontogeny of the Immune System to Promote Immune-Mediated Health for Life. Front. Immunol. 6. 10.3389/fimmu.2015.00077.

9. Halasa, N.B., Olson, S.M., Staat, M.A., Newhams, M.M., Price, A.M., Boom, J.A., Sahni, L.C., Cameron, M.A., Pannaraj, P.S., Bline, K.E., et al. (2022). Effectiveness of Maternal Vaccination with mRNA COVID-19 Vaccine During Pregnancy Against COVID-19– Associated Hospitalization in Infants Aged <6 Months — 17 States, July 2021–January 2022. MMWR Morb. Mortal. Wkly. Rep. 71, 264–270. 10.15585/mmwr.mm7107e3.

10. Halasa, N.B., Olson, S.M., Staat, M.A., Newhams, M.M., Price, A.M., Pannaraj, P.S., Boom, J.A., Sahni, L.C., Chiotos, K., Cameron, M.A., et al. (2022). Maternal Vaccination and Risk of Hospitalization for Covid-19 among Infants. N. Engl. J. Med. 387, 109–119. 10.1056/NEJMoa2204399.

11. Zerbo, O., Ray, G.T., Fireman, B., Layefsky, E., Goddard, K., Lewis, E., Ross, P., Omer, S., Greenberg, M., and Klein, N.P. (2023). Maternal SARS-CoV-2 vaccination and infant protection against SARS-CoV-2 during the first six months of life. Nat. Commun. 14, 894. 10.1038/s41467-023-36547-4.

12. Trostle, M.E., Aguero-Rosenfeld, M.E., Roman, A.S., and Lighter, J.L. (2021). High antibody levels in cord blood from pregnant women vaccinated against COVID-19. Am. J. Obstet. Gynecol. MFM 3, 100481. 10.1016/j.ajogmf.2021.100481.

13. Nir, O., Schwartz, A., Toussia-Cohen, S., Leibovitch, L., Strauss, T., Asraf, K., Doolman, R., Sharabi, S., Cohen, C., Lustig, Y., et al. (2022). Maternal-neonatal transfer of SARS-CoV-2 immunoglobulin G antibodies among parturient women treated with BNT162b2 messenger RNA vaccine during pregnancy. Am. J. Obstet. Gynecol. MFM 4, 100492. 10.1016/j.ajogmf.2021.100492.

14. Atyeo, C.G., Shook, L.L., Brigida, S., De Guzman, R.M., Demidkin, S., Muir, C., Akinwunmi, B., Baez, A.M., Sheehan, M.L., McSweeney, E., et al. (2022). Maternal immune response and placental antibody transfer after COVID-19 vaccination across trimester and platforms. Nat. Commun. 13, 3571. 10.1038/s41467-022-31169-8.

15. Atyeo, C., DeRiso, E.A., Davis, C., Bordt, E.A., De Guzman, R.M., Shook, L.L., Yonker, L.M., Fasano, A., Akinwunmi, B., Lauffenburger, D.A., et al. (2021). COVID-19 mRNA vaccines drive differential antibody Fc-functional profiles in pregnant, lactating, and nonpregnant women. Sci. Transl. Med. 13, eabi8631. 10.1126/scitranslmed.abi8631.

16. Cassidy, A.G., Li, L., Golan, Y., Gay, C., Lin, C.Y., Jigmeddagva, U., Chidboy, M.A., Ilala, M., Buarpung, S., Gonzalez, V.J., et al. (2023). Assessment of Adverse Reactions, Antibody Patterns, and 12-month Outcomes in the Mother-Infant Dyad After COVID-19 mRNA Vaccination in Pregnancy. JAMA Netw. Open 6, e2323405. 10.1001/jamanetworkopen.2023.23405.

17. Shook, L.L., Atyeo, C.G., Yonker, L.M., Fasano, A., Gray, K.J., Alter, G., and Edlow, A.G. (2022). Durability of Anti-Spike Antibodies in Infants After Maternal COVID-19 Vaccination or Natural Infection. JAMA 327, 1087. 10.1001/jama.2022.1206.

18. Zohar, T., Loos, C., Fischinger, S., Atyeo, C., Wang, C., Slein, M.D., Burke, J., Yu, J., Feldman, J., Hauser, B.M., et al. (2020). Compromised Humoral Functional Evolution Tracks with SARS-CoV-2 Mortality. Cell 183, 1508–1519.e12. 10.1016/j.cell.2020.10.052.

19. Wang, C., Li, Y., Kaplonek, P., Gentili, M., Fischinger, S., Bowman, K.A., Sade-Feldman, M., Kays, K.R., Regan, J., Flynn, J.P., et al. (2022). The Kinetics of SARS-CoV-2 Antibody Development Is Associated with Clearance of RNAemia. mBio 13, e01577–22. 10.1128/mbio.01577-22.

20. Mackin, S.R., Desai, P., Whitener, B.M., Karl, C.E., Liu, M., Baric, R.S., Edwards, D.K., Chicz, T.M., McNamara, R.P., Alter, G., et al. (2023). Fc-γR-dependent antibody effector functions are required for vaccine-mediated protection against antigen-shifted variants of SARS-CoV-2. Nat. Microbiol. 8, 569–580. 10.1038/s41564-023-01359-1.

21. Bruhns, P., and Jönsson, F. (2015). Mouse and human FcR effector functions. Immunol. Rev. 268, 25–51. 10.1111/imr.12350.

22. Atyeo, C., Pullen, K.M., Bordt, E.A., Fischinger, S., Burke, J., Michell, A., Slein, M.D., Loos, C., Shook, L.L., Boatin, A.A., et al. (2021). Compromised SARS-CoV-2-specific placental antibody transfer. Cell 184, 628–642.e10. 10.1016/j.cell.2020.12.027.

23. Jennewein, M.F., Goldfarb, I., Dolatshahi, S., Cosgrove, C., Noelette, F.J., Krykbaeva, M., Das, J., Sarkar, A., Gorman, M.J., Fischinger, S., et al. (2019). Fc Glycan-Mediated Regulation of Placental Antibody Transfer. Cell 178, 202–215.e14. 10.1016/j.cell.2019.05.044.

24. Mithal, L.B., Otero, S., Shanes, E.D., Goldstein, J.A., and Miller, E.S. (2021). Cord blood antibodies following maternal coronavirus disease 2019 vaccination during pregnancy. Am. J. Obstet. Gynecol. 225, 192–194. 10.1016/j.ajog.2021.03.035.

25. Murphy, E.A., Guzman-Cardozo, C., Sukhu, A.C., Parks, D.J., Prabhu, M., Mohammed, I., Jurkiewicz, M., Ketas, T.J., Singh, S., Canis, M., et al. (2023). SARS-CoV-2 vaccination, booster, and infection in pregnant population enhances passive immunity in neonates. Nat. Commun. 14, 4598. 10.1038/s41467-023-39989-y.

26. Prabhu, M., Murphy, E.A., Sukhu, A.C., Yee, J., Singh, S., Eng, D., Zhao, Z., Riley, L.E., and Yang, Y.J. (2021). Antibody Response to Coronavirus Disease 2019 (COVID-19) Messenger RNA Vaccination in Pregnant Women and Transplacental Passage Into Cord Blood. Obstet. Gynecol. 138, 278–280. 10.1097/AOG.0000000000004438.

27. Malek, A., Sager, R., Kuhn, P., Nicolaides, K.H., and Schneider, H. (1996). Evolution of Maternofetal Transport of Immunoglobulins During Human Pregnancy. Am. J. Reprod. Immunol. 36, 248–255. 10.1111/j.1600-0897.1996.tb00172.x.

28. Palmeira, P., Quinello, C., Silveira-Lessa, A.L., Zago, C.A., and Carneiro-Sampaio, M. (2012). IgG Placental Transfer in Healthy and Pathological Pregnancies. Clin. Dev. Immunol. 2012, 1–13. 10.1155/2012/985646.

29. Clements, T., Rice, T.F., Vamvakas, G., Barnett, S., Barnes, M., Donaldson, B., Jones, C.E., Kampmann, B., and Holder, B. (2020). Update on Transplacental Transfer of IgG Subclasses: Impact of Maternal and Fetal Factors. Front. Immunol. 11, 1920. 10.3389/fimmu.2020.01920.

30. Bartsch, Y.C. (2022). Preserved recognition of Omicron spike following COVID-19 messenger RNA vaccination in pregnancy. Am. J. Obstet. Gynecol. 227, 493.e1–493.e7.

31. Ou, J., Lan, W., Wu, X., Zhao, T., Duan, B., Yang, P., Ren, Y., Quan, L., Zhao, W., Seto, D., et al. (2022). Tracking SARS-CoV-2 Omicron diverse spike gene mutations identifies multiple inter-variant recombination events. Signal Transduct. Target. Ther. 7, 138. 10.1038/s41392-022-00992-2.

32. Bartsch, Y.C., St. Denis, K.J., Kaplonek, P., Kang, J., Lam, E.C., Burns, M.D., Farkas, E.J., Davis, J.P., Boribong, B.P., Edlow, A.G., et al. (2022). SARS-CoV-2 mRNA vaccination elicits robust antibody responses in children. Sci. Transl. Med. 14, eabn9237. 10.1126/scitranslmed.abn9237.

33. Yang, Y.J., Murphy, E.A., Singh, S., Sukhu, A.C., Wolfe, I., Adurty, S., Eng, D., Yee, J., Mohammed, I., Zhao, Z., et al. (2022). Association of Gestational Age at Coronavirus Disease 2019 (COVID-19) Vaccination, History of Severe Acute Respiratory Syndrome Coronavirus 2 (SARS-CoV-2) Infection, and a Vaccine Booster Dose With Maternal and Umbilical Cord Antibody Levels at Delivery. Obstet. Gynecol. 139.

34. Gray, K.J., Bordt, E.A., Atyeo, C., Deriso, E., Akinwunmi, B., Young, N., Baez, A.M., Shook, L.L., Cvrk, D., James, K., et al. (2021). Coronavirus disease 2019 vaccine response in pregnant and lactating women: a cohort study. Am. J. Obstet. Gynecol. 225, 303.e1–303.e17. 10.1016/j.ajog.2021.03.023.

35. Mor, G., and Cardenas, I. (2010). The Immune System in Pregnancy: A Unique Complexity. Am. J. Reprod. Immunol. 63, 425–433. 10.1111/j.1600-0897.2010.00836.x.

36. Irrgang, P., Gerling, J., Kocher, K., Lapuente, D., Steininger, P., Habenicht, K., Wytopil, M., Beileke, S., Schäfer, S., Zhong, J., et al. (2023). Class switch toward noninflammatory, spike-specific IgG4 antibodies after repeated SARS-CoV-2 mRNA vaccination. Sci. Immunol. 8, eade2798. 10.1126/sciimmunol.ade2798.

37. Korobova, Z.R., Zueva, E.V., Arsentieva, N.A., Batsunov, O.K., Liubimova, N.E., Khamitova, I.V., Kuznetsova, R.N., Rubinstein, A.A., Savin, T.V., Stanevich, O.V., et al. (2022). Changes in Anti-SARS-CoV-2 IgG Subclasses over Time and in Association with Disease Severity. Viruses 14, 941. 10.3390/v14050941.

38. SMFM (2023). RSV Vaccination in Pregnancy (Society for Maternal Fetal Medicine).

39. Kampmann, B., Madhi, S.A., Munjal, I., Simões, E.A.F., Pahud, B.A., Llapur, C., Baker, J., Pérez Marc, G., Radley, D., Shittu, E., et al. (2023). Bivalent Prefusion F Vaccine in Pregnancy to Prevent RSV Illness in Infants. N. Engl. J. Med. 388, 1451–1464. 10.1056/NEJMoa2216480.

40. Castro-Dopico, T., and Clatworthy, M.R. (2019). IgG and Fcγ Receptors in Intestinal Immunity and Inflammation. Front. Immunol. 10, 805. 10.3389/fimmu.2019.00805.

41. Bournazos, S., Gupta, A., and Ravetch, J.V. (2020). The role of IgG Fc receptors in antibody-dependent enhancement. Nat. Rev. Immunol. 20, 633–643. 10.1038/s41577-020-00410-0.

42. Lee, Y.-C., and Lin, S.-J. (2013). Neonatal Natural Killer Cell Function: Relevance to Antiviral Immune Defense. Clin. Dev. Immunol. 2013, 1–6. 10.1155/2013/427696.

43. Atyeo, C., Shook, L.L., Nziza, N., Deriso, E.A., Muir, C., Baez, A.M., Lima, R.S., Demidkin, S., Brigida, S., De Guzman, R.M., et al. (2023). COVID-19 booster dose induces robust antibody response in pregnant, lactating, and nonpregnant women. Am. J. Obstet. Gynecol. 228, 68.e1–68.e12. 10.1016/j.ajog.2022.07.014.

44. Pillai, S. (2023). Is it bad, is it good, or is IgG4 just misunderstood? Sci. Immunol. 8, eadg7327. 10.1126/sciimmunol.adg7327.

45. Uversky, V.N., Redwan, E.M., Makis, W., and Rubio-Casillas, A. (2023). IgG4 Antibodies Induced by Repeated Vaccination May Generate Immune Tolerance to the SARS-CoV-2 Spike Protein. Vaccines 11, 991. 10.3390/vaccines11050991.

46. FDA (2023). FDA Approves First Vaccine for Pregnant Individuals to Prevent RSV in Infants (Food and Drug Administration).

47. CDC (2023). CDC recommends new vaccine to help protect babies against severe respiratory syncytial virus (RSV) illness after birth (US Department of Health and Human Services, CDC).

48. ACOG (2023). ACOG Unequivocally Supports ACIP’s Recommendation Approving Use of Maternal RSV Vaccine in Pregnancy (American College of Obstetricians and Gynecologists).

49. Bordt, E.A., Shook, L.L., Atyeo, C., Pullen, K.M., De Guzman, R.M., Meinsohn, M.-C., Chauvin, M., Fischinger, S., Yockey, L.J., James, K., et al. (2021). Maternal SARS-CoV-2 infection elicits sexually dimorphic placental immune responses. Sci. Transl. Med. 13, eabi7428. 10.1126/scitranslmed.abi7428.

50. Lipschuetz, M., Guedalia, J., Cohen, S.M., Sompolinsky, Y., Shefer, G., Melul, E., Ergaz-Shaltiel, Z., Goldman-Wohl, D., Yagel, S., Calderon-Margalit, R., et al. (2023). Maternal third dose of BNT162b2 mRNA vaccine and risk of infant COVID-19 hospitalization. Nat. Med. 29, 1155–1163. 10.1038/s41591-023-02270-2.

51. Brown, E.P., Licht, A.F., Dugast, A.-S., Choi, I., Bailey-Kellogg, C., Alter, G., and Ackerman, M.E. (2012). High-throughput, multiplexed IgG subclassing of antigen-specific antibodies from clinical samples. J. Immunol. Methods 386, 117–123. 10.1016/j.jim.2012.09.007.

52. McInnes, L., Healy, J., and Melville, J. (2020). UMAP: Uniform Manifold Approximation and Projection for Dimension Reduction. Preprint at arXiv.

53. Wold, S., Esbensen, K., and Geladi, P. Principal Component Analysis.

54. Tibshirani, R. (1996). Regression Shrinkage and Selection Via the Lasso. J. R. Stat. Soc. Ser. B Methodol. 58, 267–288. 10.1111/j.2517-6161.1996.tb02080.x.

